# Treatment of obstructive sleep apnea using CPAP and cognitive impairment

**DOI:** 10.1101/2025.08.07.25333263

**Authors:** Anastasya Maria Kosasih, Juliana Tereza Colpani, Laureen YT Wang, Serene Wong, Yi-Hui Ou, Chi-Hang Lee

## Abstract

**Study objective:** To evaluate the association between obstructive sleep apnea (OSA) and cognitive impairment by comparing individuals with continuous positive airway pressure (CPAP)-treated OSA, untreated OSA, and those without OSA.

**Methods:** In this cross-sectional study, participants were divided into 3 groups based on overnight sleep study and CPAP use: (1) OSA–CPAP group, (2) OSA–no treatment group, and (3)non–OSA group. The primary outcome was the prevalence of cognitive impairment among 3 groups measured by Montreal Cognitive Assessment (MoCA) and the secondary outcome was ambulatory blood pressure monitoring (ABPM).

**Results:** A total of 113 participants (male 69.9%, median age: 57.0 years) are enrolled, comprising 50 participants in the OSA–CPAP group, 50 participants in the OSA–no treatment group, and 13 participants in the non–OSA group. The cognitive impairment prevalence was 46.0% in the OSA–CPAP group, 70.0% in the OSA–no treatment group, and 46.0% in the non-OSA groups (p=0.038). Subdomain analysis showed significantly higher scores in memory and visuospatial abilities in the OSA–CPAP and the non–OSA group (p=0.039 and p=0.005). Post-hoc analysis revealed significantly higher MoCA scores and lower prevalence of cognitive impairment in the OSA–CPAP group compared to OSA–no treatment group (p=0.002 and p=0.015). No significant differences in ABPM parameters were observed.

**Conclusion:** CPAP therapy in patients with OSA is associated with better cognitive performance and lower prevalence of cognitive impairment. These findings suggest a potential role for OSA treatment preserving cognitive function, however further longitudinal studies are needed.

**BRIEF SUMMARY:** The relationship between obstructive sleep apnea (OSA) and cognitive impairment remains controversial. Most prior studies relied on retrospective analyses or selected participants using strict inclusion and exclusion criteria, limiting their real-world applicability. In this study, we analyzed data from unselected patients attending a sleep clinic. We found that patients with treated OSA had a similar prevalence of cognitive impairment as those without OSA, and both groups had a lower prevalence than patients with untreated OSA.

## INTRODUCTION

Cognitive impairment refers to measurable declines in key mental functions such as memory, attention, learning, and decision-making.^1^ It is increasingly common in aging populations and becomes more prevalent with advancing age.^2^ It spans a spectrum, with mild cognitive impairment (MCI) representing an intermediate stage between normal aging and dementia. While some individuals with MCI may stabilize or improve, others progress to irreversible cognitive decline.^1^ Cognitive impairment can seriously affect daily life by diminishing memory, concentration, and decision-making abilities ─ these frequently lead to challenges in employment and everyday tasks, reducing independence and quality of life, and increasing caregiver burden.^3^

Sleep is essential for brain health, yet sleep disorders are common worldwide. Obstructive sleep apnea (OSA) is a chronic condition marked by repeated upper airway obstruction during sleep, leading to recurrent hypoxia, arousal, and blood pressure surge during sleep.^4^ Patients with untreated OSA frequently experience excessive daytime sleepiness and impaired mental function.^5–7^ Intuitively, patients with OSA, especially untreated, may be a risk factor for cognitive impairment. However, data in the literature on between OSA and cognitive impairment have been conflicting, with observational studies showing an association between OSA and cognitive impairment,^1,8^ but studies on the effects of OSA treatment on cognitive function have produced inconsistent results.^9–12^

We conducted a cross-sectional study in real-world sleep clinic settings to assess cognitive function in three groups: individuals with OSA treated with continuous positive airway pressure (CPAP) for at least six months, individuals with untreated OSA, and those without OSA. Cognitive performance was evaluated using the Montreal Cognitive Assessment (MoCA),^13^ a validated tool for early detection of cognitive impairment. We hypothesized that compared with untreated OSA, treatment of OSA with CPAP is associated with lower prevalence of cognitive impairment.

## MATERIALS AND METHODS

### Study Design

This investigator-initiated, cross-sectional study was conducted at the outpatient sleep clinics of two public hospitals in Singapore. The aim was to examine the relationship between OSA, CPAP therapy, and the prevalence of MCI (ClinicalTrials.gov identifier: NCT06463002). The study was funded by a competitive public research grant from Singapore’s Ministry of Health. Adult patients attending the sleep clinics for follow-up visits and who had undergone sleep study were invited to participate in a cognitive assessment. Assessments were conducted by an investigator who is a medically trained physician certified in administering the MoCA questionnaire. Exclusion criteria included individuals with known OSA who were using non-CPAP treatments—such as mandibular advancement devices, upper airway surgery, or hypoglossal nerve stimulation—as well as those with a history of heart failure, atrial fibrillation, acute coronary syndrome within the past three months, dementia, or prior stroke.

Baseline characteristics collected included age, body mass index (BMI), past medical history, education level, neck, waist, and hip circumference, and Epworth Sleepiness Scale (ESS) scores. Participants were categorized into three groups based on their overnight sleep study results and CPAP usage:

1. **OSA–CPAP group**: Apnea Hypopnea Index (AHI) ≥15 events/hour and on CPAP therapy for at least six months.
2. **OSA–no treatment group**: AHI ≥15 events/hour and had not used CPAP or any other approved OSA treatment for at least six months prior to the clinic visit.
3. **Non-OSA group**: AHI <15 events/hour and had not used CPAP or any other approved OSA treatment for at least six months prior to the clinic visit.

### Ethics statement

The study was approved by the institutional review board (Domain Specific Review Board-C: 2023/01017). All participants provided written informed consent in the presence of a witness. Participants were recruited between July 4, 2024, and April 16, 2025.

### Measurements and Outcomes

The primary outcome of this study was the prevalence of cognitive impairment among 3 groups of patients. Cognitive function was assessed using the MoCA,^14^ a widely used screening tool designed to detect cognitive impairment and early signs of dementia. The MoCA evaluates multiple cognitive domains, including attention, memory, language, visuospatial abilities, executive function, and orientation. It consists of a series of standardized tasks and questions targeting various aspects of cognition. In this study, the digital version of the MoCA was administered by trained healthcare professionals. The tool has been validated in diverse populations and is known for its high sensitivity in detecting cognitive impairment. Participants were classified as having MCI based on their MoCA scores, adjusted for educational level, according to a validation study conducted in local population.^15^ Specifically, a MoCA score of <27 was considered indicative of cognitive impairment in participants with >10 years of formal education, while a score of <26 was used for those with ≤ 10 years of education.^15^

An additional (optional) secondary outcome was ambulatory blood pressure monitoring (ABPM), a widely accepted non-invasive method to evaluate 24-hour blood pressure control. Ambulatory BP monitoring will be performed over 24 hours using a clinically approved oscillometric device (Welch Allyn ABPM 7100, Welch Allyn, Skaneateles Falls, New York, USA) programmed to record BP every 30 min. Participants were asked to complete a diary card and use an event marker to indicate sleep and wake periods. ABPM data were considered valid and included in the analysis if the device recorded a minimum of 21 readings within the 24-hour monitoring period.

### Statistical Analysis

Descriptive statistics were used to summarize the baseline characteristics of the participants. Continuous variables were presented as mean ± standard deviation (SD) or median with interquartile range (IQR), depending on the distribution, while categorical variables were reported as frequencies and percentages. Exploratory analyses were conducted using the chi-square test for categorical variables and the Kruskal Wallis test for continuous variables and Dunn Pairwise comparison and two-proportion Z test as post-hoc analysis to identify overall differences between multiple groups. Box plots were used to visually compare MoCA scores, 24-hour mean arterial pressure, and both daytime and nighttime blood pressure across the three study groups. All statistical analyses were performed using Stata BE version 18.0 (Stata Corp).

## RESULTS

### Baseline Characteristics

Between July 2024 and April 2025, a total of 113 participants (male 69.9%, n=79; age 57.0 [44.0 - 66.0] years) were enrolled into this study. The median BMI of was 27.2 (IQR, 24.5-31.4) kg/m^2^. Most of the participants (88.5%, 100/113) have received at least 10 years of formal education. Details of the demographic and clinical characteristics of the study participants are shown in **Table 1**.

There were 50 participants in the OSA - CPAP group. All these participants have been using CPAP, for a median duration of 28.0 (18.0 – 33.0) months. The 95^th^ pressure was 10.1 (8.4 – 12.4) cmH_2_O. The average CPAP usage was 5.7 ± 1.6 hours/night, with 68.7 ± 25.5% of the participants used at least 4 hours/night. The residual AHI was 1.7 (1.1 – 2.8) hour/events. Besides, there were 50 participants in the OSA - no treatment group, and 13 participants in the non - OSA group. None of the patients in these latter 2 groups were using CPAP or other guideline-approved OSA therapy.

Regarding cardiovascular comorbidities, 51.3% (58/113) have hypertension, and among these 45.0% (27/58) had a duration of hypertension for >10 years. No significant differences were observed in the use of antihypertensives medications among the three groups. For OSA diagnostic assessment. 81.6% (84/113) of the participants underwent an American Academy of Sleep Medicine (AASM) level 1 polysomnography, while the remainder underwent home-based sleep study using level 3 portable diagnostic device. Additionally, 22.9% (26/113) of participants exhibited excessive day time sleepiness, as indicated by ESS score >10 points **(Table 3)**. Details of the sleep study results are summarized in **Table 2**.

### MoCA

The detailed results of the MoCA score are shown in **Table 3**. Overall, the prevalence of mild cognitive impairment (based on the aforementioned definition) differed among the 3 groups: 46.0% (23/50) in the OSA - CPAP group, 70.0% (35/50) in the OSA - no treatment group, and 46.0% (6/13) in the non - OSA group (p=0.038). As an exploratory sensitivity analysis, the raw MoCA scores without taking the years of education into consideration also differed among the 3 groups (p=0.005).

Analysis of the MoCA subdomains revealed significant differences in memory performance and visuospatial abilities, specifically in tasks involving trail making, clock drawing, and cube copying. Participants in the OSA - CPAP group and the non - OSA group achieved significantly higher (better) scores in memory and visuospatial abilities cognitive domains compared to those in the OSA without treatment group (p=0.039 and p=0.005) **(Table 3)**.

Post-hoc analysis using Dunn’s pairwise comparison reveals those participants in the OSA - CPAP group had significantly higher (better) MoCA scores compared to those in the OSA - no CPAP group (p=0.002) **(Figure 1)**. However, no significant differences in MoCA scores were found between the OSA - CPAP group and the non - OSA group (p=0.566), or between the OSA-no CPAP group and the non-OSA group (p=0.330).

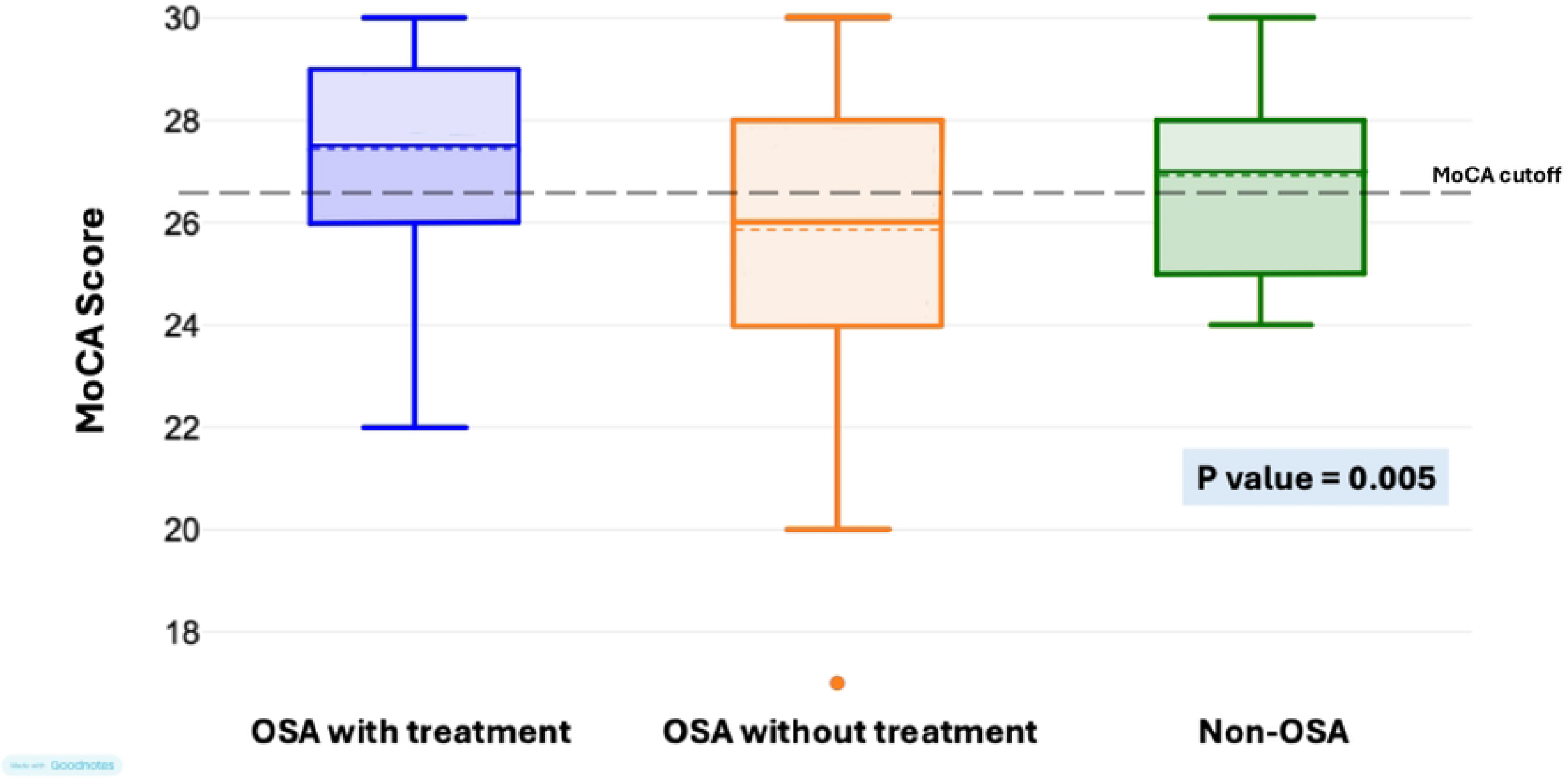

A two-proportion Z-test for cognitive impairment revealed lower prevalence of cognitive impairment in participants in the OSA - CPAP group compared to OSA - no treatment group (diff= -24%; 95% CI: -42.8% to -5.2%; p=0.015). In contrast, there was no significant difference in prevalence of cognitive impairment between patients in the OSA-CPAP group and non-OSA group (diff= 0%; 95% CI: -30.4% to 30.4%; p=1.000). While the cognitive impairment prevalence in OSA-no-CPAP group is higher compared to non-OSA group, this difference did not show statistical significance (diff= 24%; 95% CI: -5.9% to 53.9%; p=0.106).

### Ambulatory blood pressure

A total of 17 participants underwent ABPM, comprising 8 individuals from the OSA– CPAP group, 7 individuals from OSA-no treatment group, and 2 individuals from the non-OSA group. No significant differences were observed among the three groups with respect to 24-hour mean BP, daytime and nighttime blood pressure measurements, systolic BP dipping, or pulse pressure parameters **(Table 4)**.

## DISCUSSION

In this cross-sectional study involving real-world sleep clinic patients, we found that patients with OSA (AHI ≥15 events/hour) who are treated with CPAP and patients without OSA (AHI <15 events/hour) have a lower prevalence of mild cognitive impairment than patients with untreated OSA. Furthermore, OSA–CPAP patients demonstrated better performance in memory and visuospatial cognitive subdomains of the MoCA test. No significant differences were observed in ABPM parameters among the study groups, which may be due to limited number of participants. These findings provide evidence that OSA is associated with mild cognitive impairment, and treatment of OSA with CPAP may potentially help preserve cognitive function. Incorporating cognitive screening, especially in elderly, may assist in identifying for neurocognitive decline and inform future research and intervention strategies.

Previous studies have demonstrated that OSA contributes to cognitive decline through repeated episodes of intermittent hypoxia and sleep fragmentation. The significant cognitive benefit observed in OSA–CPAP group, aligns with earlier studies suggesting partial reversibility of neurocognitive treatment following the effective treatment of OSA. Specifically, CPAP therapy has been shown to improve working memory, processing speed, and attention span, with the benefit observable in the first 3 months of the treatment. Our study extends these findings by demonstrating the statically significant improvement in MoCA subdomains score, memory and visuospatial cognition among the OSA–group. In addition, the mechanisms linking OSA to cognitive decline are multifactorial and likely involve chronic intermittent hypoxia, oxidative stress, systemic inflammation, endothelial dysfunction, and cerebrovascular dysregulation. Recurrent desaturation episodes lead to blood–brain barrier disruption and neuroinflammation, which can damage neurons in key cognitive areas such as the hippocampus and prefrontal cortex.^16^ CPAP therapy mitigates these deleterious effects by restoring nocturnal oxygenation, reducing arousal frequency, and improving slow-wave sleep quality—factors that are essential for memory consolidation and neural repair. The specific improvements observed in visuospatial and memory domains in this study may reflect the selective vulnerability of parietal and temporal brain regions to chronic intermittent hypoxia. These findings suggest that timely and sustained CPAP use may slow the progression of early neurocognitive changes in patients with OSA.

Intuitively, patients with cognitive impairment are believed to have lower adherence to CPAP for OSA due to factors such as difficulty operating the equipment, forgetting to use the device, lack of insight into their condition, and behavioral issues that interfere with nightly use. To date, no study has prospectively examined predictors of CPAP adherence in a well-characterized, large sample of OSA patients with cognitive impairment, whose memory deficits may affect their ability to use CPAP. The *Memories 2* study—a large, prospective investigation—reported predictors of CPAP adherence in older adults with amnestic mild cognitive impairment and moderate to severe OSA.^17^ The result shows that, among individuals at high risk for progression to dementia due to Alzheimer’s disease, 75% of participants adhered to CPAP for over 3 months, using the standard threshold of at least 4 hours per night. The average CPAP usage was 5.15 ± 2.50 hours per night. Notably, there was no significant association between CPAP adherence and the severity of cognitive impairment. Although further research is needed, current evidence suggests that older adults with mild cognitive impairment and mild dementia are cognitively capable of using CPAP. Therefore, cognitive impairment alone should not be considered a barrier to prescribing CPAP therapy.

Several limitations warrant consideration in this study. We set out to recruit 50 participants for each of the 3 groups, we reached the recruitment target for both the OSA – CPAP group and OSA – no treatment group. For the non-OSA group, we were only able to recruit 13 participants. This is because there is a high prevalence of OSA in Singapore.^18^ Due to the relatively low level of awareness,^19^ patients without OSA are frequent asymptomatic and hence would not have been referred for sleep study. Patients who adhere to CPAP therapy may also be more likely to engage in other healthy lifestyle behaviors, such as maintaining a balanced diet or exercising regularly, which could contribute to the lower prevalence of cognitive impairment observed in the OSA–CPAP group. This study aimed to study the effect of CPAP, although there are other effective OSA therapies, such as mandibular advancement device.^20^ The cross-sectional design also limits our ability to track longitudinal changes in cognitive function. Further studies should focus on longitudinal, multi-center trials with larger sample sizes to validate the cognitive benefits of CPAP over time. The integration of neuroimaging modalities and neuroinflammation biomarkers could elucidate structural and functional brain change, and mechanistic pathways in response to treatment.

## Conclusion

We found that patients with OSA who use CPAP therapy regularly have a lower prevalence of cognitive impairment than those who do not use CPAP. The prevalence of cognitive impairment in patients with OSA who use CPAP therapy regularly is similar to those without OSA. While these findings support the importance of early diagnosis and treatment adherence, further longitudinal studies are needed. Routine cognitive screening may help identify at-risk individuals and inform for future research.

## Data Availability

All relevant data are within the manuscript and its Supporting Information files.

## ABBREVIATIONS

ABPM: ambulatory blood pressure monitoring
AHI: apnea hypopnea index
CPAP: continuous positive airway pressure
IQR: interquartile range
MCI: mild cognitive impairment
MoCA: Montreal Cognitive Assessment
OSA: Obstructive sleep apnea
SD: standard deviation

